# From Theory to Practice: A Scoping Review of Theory-Based Educational Approaches in Hemodialysis Care

**DOI:** 10.1101/2025.09.11.25335569

**Authors:** Atikah Fatmawati, Moses Glorino Rumambo Pandin, Anndy Prastya, Fitria Wahyu Ariyanti

## Abstract

**Background:** Patients on hemodialysis face complex challenges in treatment adherence, self-care, and quality of life. Patient education is central to overcoming these challenges, yet many interventions are implemented without a theoretical foundation, potentially limiting their effectiveness. Examining theory-based approaches may provide valuable insights for designing more impactful educational strategies.

**Objective:** This scoping review aimed to map theoretical frameworks underpinning patient education interventions in hemodialysis care and to explore their reported outcomes.

**Methods:** Following the Arksey and O’Malley framework and reported according to PRISMA-ScR, a systematic search was conducted in PubMed, Scopus, CINAHL, and Web of Science for studies published between 2020 and 2025. Eligible studies included educational interventions for adult hemodialysis patients that explicitly applied a theoretical or conceptual framework. Data were extracted on study design, population, educational approach, theoretical framework, and outcomes.

**Results:** A limited but growing body of research incorporated theory-based educational approaches in hemodialysis care. Frequently applied frameworks included the Health Belief Model, Self-Determination Theory, 5A Self-Management Model, and the Temporal Self-Regulation Theory (TST). Interventions grounded in theory demonstrated improvements in treatment adherence, self-care behaviors, self-efficacy, and quality of life. However, heterogeneity in theoretical application and inconsistent reporting were noted across studies.

**Conclusion:** Theory-based educational interventions show promising benefits for hemodialysis patients, particularly in supporting adherence and self-management. Nonetheless, the limited number of studies highlights the need for more robust, theory-driven research to guide best practices in patient education and optimize patient outcomes.

## Introduction

Chronic kidney disease (CKD) is a global health problem with a growing burden year after year. An estimated 3.7–4 million people worldwide receive kidney replacement therapy (KRT), and hemodialysis is the most widely used modality, surpassing peritoneal dialysis and kidney transplantation (Liang, Lin, & Wu, 2022; International Society of Nephrology (ISN), 2023). This trend aligns with the increasing incidence of non-communicable diseases, particularly diabetes mellitus and hypertension, which are major risk factors for CKD.

Hemodialysis is a life-sustaining therapy for patients with end-stage renal disease, but this procedure poses complex challenges that extend beyond the dialysis sessions themselves. Patients are required to adhere to strict fluid and dietary restrictions, maintain vascular access, and consistently practice self-care practices. Non-adherence to the regimen is associated with increased morbidity, higher frequency of hospitalizations, and reduced quality of life (Friedrich et al., 2025; Sajjadi, Ghafourifard, & Tayebi Khosroshahi, 2024).

In Indonesia, the burden of hemodialysis patients has increased significantly. Data from the Indonesian Renal Registry (IRR) reports a surge in the number of active patients and new cases of hemodialysis in the past decade, with a more than tenfold increase from 2011 to 2021 (Indonesian Renal Registry, 2022). International comparative analysis places Indonesia as one of the countries with the fastest increase in dialysis prevalence in the world (United States Renal Data System [USRDS], 2022). This situation underscores the urgency of developing effective educational interventions to improve adherence, quality of life, and clinical outcomes for hemodialysis patients.

Patient education is recognized as a key pillar for improving adherence and supporting self-management in individuals undergoing hemodialysis. However, many educational interventions still focus solely on information delivery without a clear theoretical framework (Liu et al., 2024; van Eck van der Sluijs et al., 2021). The lack of a theoretical foundation makes it difficult to explain the mechanisms of behavior change, thus limiting the effectiveness and replication of educational interventions across healthcare contexts.

In recent years, scientific evidence has demonstrated the benefits of theory-based educational approaches for hemodialysis patients. Educational programs utilizing Self-Determination Theory (SDT) have been shown to improve knowledge, self-management skills, and psychosocial outcomes such as reduced anxiety and depression (Zhang et al., 2025). Similarly, interventions using the Health Belief Model (HBM) through visualization-based health education can improve adherence behavior, while dietary programs based on self-efficacy theory are effective in improving dietary adherence and patient clinical outcomes (Peng et al., 2025). Behavioral learning strategies such as the teach-back method are also effective in increasing knowledge retention and self-efficacy (Xia et al., 2024).

Despite these promising findings, the application of theoretical models in hemodialysis education remains inconsistent. Many studies do not explicitly report the theoretical framework used, the outcomes measured vary widely, and long-term evaluation is limited (Sajjadi et al., 2024; van Eck van der Sluijs et al., 2021). This situation indicates a knowledge gap regarding which theories are most effective, how they are applied, and the specific outcomes they influence, including adherence, quality of life, and clinical parameters.

Therefore, a scoping review focusing on theory-based educational models in hemodialysis patient care is crucial and urgent. This review is expected to map the theoretical frameworks used, the educational strategies implemented, and the intended outcomes, thereby providing a strong foundation for designing more effective, evidence-based educational interventions in the future.

## Method

### Data Retrieval Strategy

This study is a scoping review to systematically answer the research question and is structured according to the Preferred Reporting Items for Systematic Reviews and Meta-Analyses (PRISMA) 2020 guidelines (Page et al., 2021). This study reviews the theories used to inform health education for hemodialysis patients. The detailed activities include determining data search strategies and/or information sources, selecting studies through quality assessment based on eligibility criteria and quality assessment instruments, and then using data synthesis and extraction.

The research question of this scoping review is “What theories have been used to inform educational interventions for hemodialysis patients, how are they implemented, and what outcomes are reported?” The literature search was conducted in five international databases: CINAHL (Ebsco), Scopus, Web of Science, PubMed, and ProQuest. The keywords and Boolean operators used in the literature search were “hemodialysis” OR “dialysis” AND “patient education” OR “health education” OR “self-management education” AND “theory” OR “model” OR “framework”.

### Eligibility Criteria

Eligibility criteria in this study included inclusion and exclusion criteria. The inclusion criteria in this study were studies reporting educational interventions in adult hemodialysis patients based on behavioral theories or models, all types of research designs (RCTs, quasi-experimental, pre-post, and observational studies), articles published between 2020 and 2025, and articles in English. Meanwhile, the exclusion criteria were studies that did not mention behavioral theories or models as the basis for education, pediatric populations (<18 years), or non-hemodialysis kidney failure patients, review articles, editorials, commentaries, or case reports. In addition, to limit the scope of the study, the researchers used the PCC (Population, Concept, Context) method, as shown in the following table:

### Quality Assessment

Literature selection used the PRISMA (Preferred Reporting Items for Systematic Reviews and Meta-analyses) method. The PRISMA Flow Diagram in this study is shown in Figure 1.

**Figure 1.**
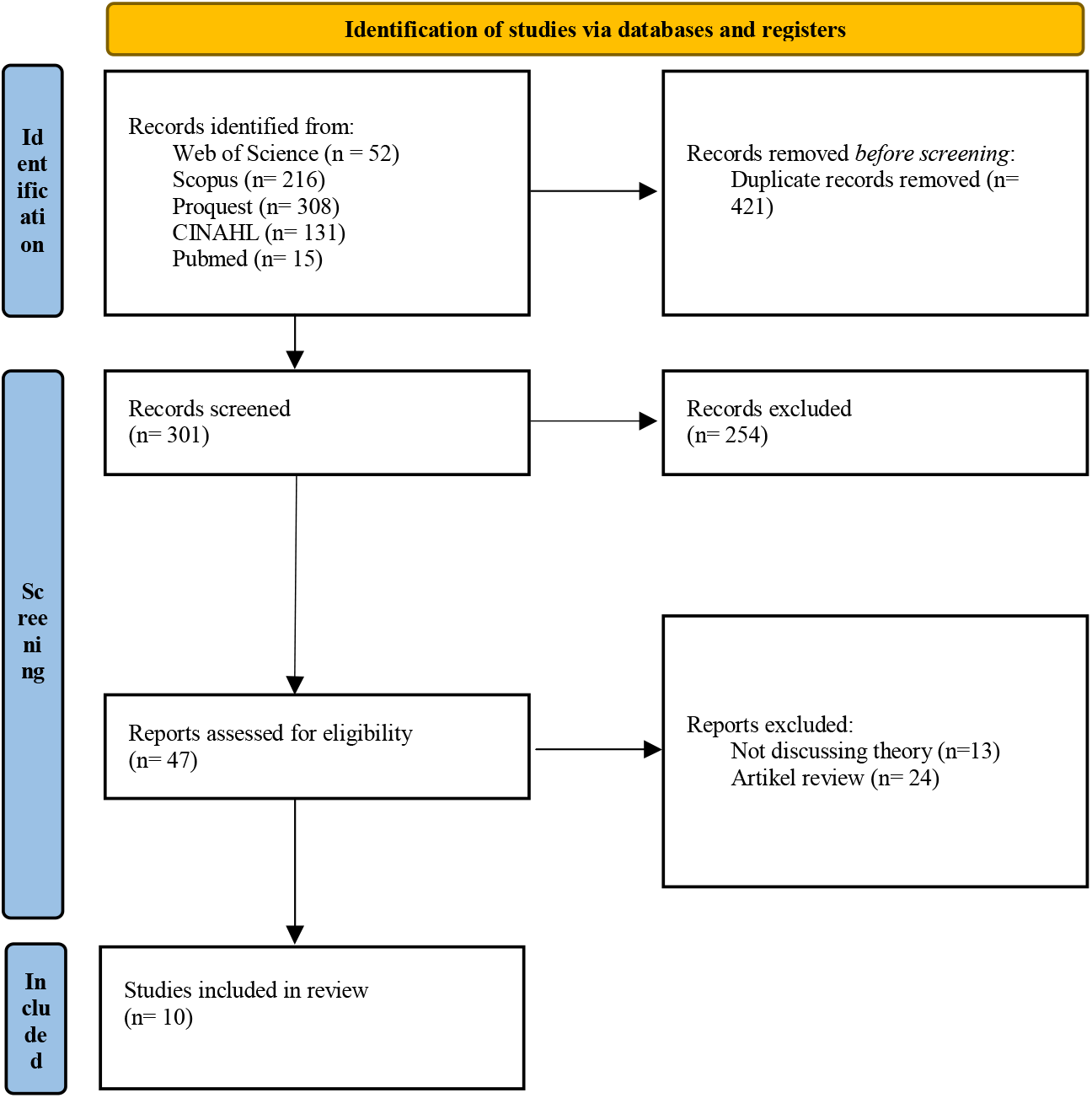
PRISMA *Flow Diagram*.

### Synthesis Data

The data synthesis process in this study was conducted by comparing literature that met quality assessments and inclusion and exclusion criteria. The data synthesis focused on the research objective of analyzing theories used to inform educational interventions for hemodialysis patients.

### Data Extraction

The data extraction output is in the form of a table consisting of the title, researcher’s name and year of publication, study design, participants, theory/theoretical framework, measured outcomes, and main findings.

## Results

Based on the selection process, a scoping review of the theories used to underlie educational interventions for hemodialysis patients identified 10 articles for further analysis. A summary of the article analysis of the theories used to underlie educational interventions for hemodialysis patients is presented in Table 2.

**Table 1.** PCC Summary.

| Componen | Information | Detail |
| --- | --- | --- |
| *Population* (P) | Adult hemodialysis patients (≥18 years) receiving regular hemodialysis therapy | Patients in hospitals, clinics, or dialysis centers; includes all medical conditions related to CKD/ESKD; no geographic limitations |
| *Concept* (C) | Educational interventions based on theory or behavioral models | Self-Determination Theory (SDT), Health Belief Model (HBM) , Social Cognitive Theory (SCT), Self-Efficacy, Teach-back, Visualisation/Multimedia Education, Multimodal Education |
| *Context* (C) | Hemodialysis services | All hemodialysis service settings: hospitals, clinics, dialysis centers; interventions can be face-to-face, group, digital, or a combination; include patient-nurse/doctor/educator interactions |

**Table 2.**
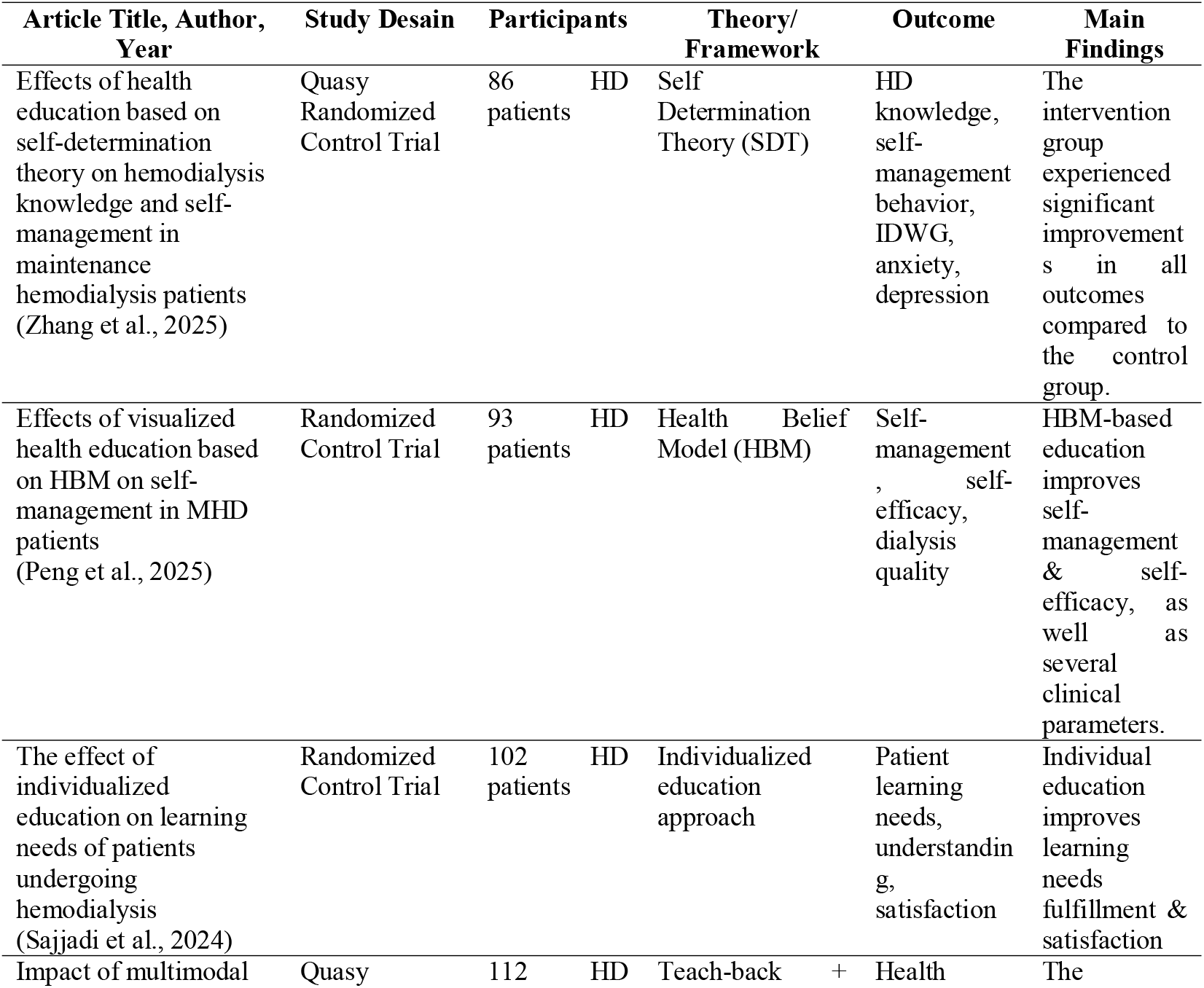
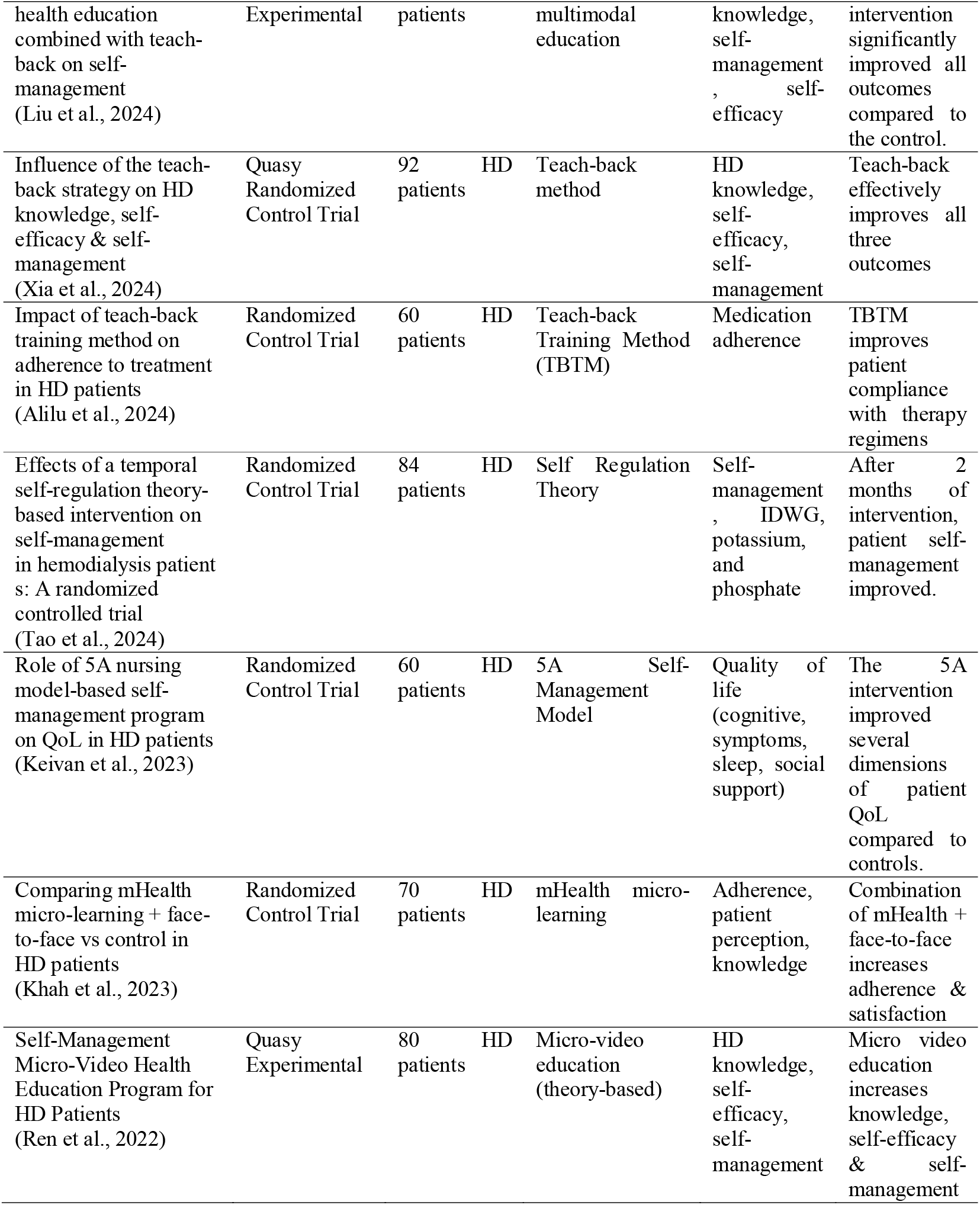
Data Extraction Results.

## Discussion

A review of nine articles on hemodialysis patient education revealed a diversity of approaches to intervention design, particularly regarding the use of theoretical frameworks. Three studies explicitly used formal theory as the basis for the intervention. Zhang et al. (2025) applied Self-Determination Theory (SDT) in an educational program, which was shown to improve patient knowledge and self-management skills, as well as reduce anxiety and depression. These findings align with evidence that SDT can foster intrinsic motivation and sustainable health behaviors. The use of a clear theory allows for scientific explanation of the mechanisms of behavior change, while also strengthening the program’s potential for replication in different clinical contexts.

Furthermore, Peng et al. (2025) used the Health Belief Model (HBM) through a visualization-based educational approach. This study reported significant improvements in self-management, self-efficacy, and dialysis quality. The HBM framework helps explain how individual perceptions of vulnerabilities and benefits influence adherence behavior. The application of the HBM in hemodialysis education shows great potential, particularly because the theory emphasizes changes in perceived risks and barriers that often hinder patients from adhering to therapy regimens. Similarly, Keivan et al. (2023) explicitly applied the 5A Self-Management Model in education, resulting in improved quality of life, particularly in physical symptoms, cognitive aspects, and social support. This model emphasizes assessment, advice, agreement, assistance, and arrangement, enabling patients to actively participate in their disease management.

On the other hand, the majority of other articles rely on an implicit educational approach without a formal theoretical framework. Sajjadi et al. (2024) developed individualized education based on patient needs, which successfully increased patient satisfaction and perceived learning needs, but without an explicit theoretical basis. Liu et al. (2024) and Xia et al. (2024) utilized a teach-back strategy to improve knowledge retention, self-efficacy, and self-management. While effective, this approach focuses more on pedagogical methods than on the psychological mechanisms that explain why behavioral changes are sustained long-term.

Alilu et al. (2024) reported the effectiveness of teach-back in improving patient adherence to hemodialysis regimens, while Khah et al. (2023) demonstrated that the use of micro-learning-based mHealth combined with face-to-face interactions can improve patient adherence, knowledge, and satisfaction. These studies confirm that innovative technology-based strategies and interactive communication have potential, but the lack of a theoretical framework makes the results difficult to generalize. Similarly, Ren et al. (2022) demonstrated the effectiveness of educational micro-videos in improving knowledge and self-efficacy, but again, the psychosocial mechanisms underlying the intervention’s success were not theoretically addressed.

The dominant trend toward implicit educational interventions indicates that most studies focus more on innovative delivery formats than on integrating theoretical frameworks. This creates limitations in explaining the mechanisms of behavior change. Without a theoretical foundation, it is difficult to ensure that interventions can be consistently replicated across different populations or healthcare systems. Therefore, the effectiveness achieved in these studies is contextual and at risk of being unsustainable in the long term.

A recent study by Tao et al. (2024) adds an important perspective by testing an educational intervention based on Temporal Self-Regulation Theory (TST) in hemodialysis patients through a randomized controlled trial. TST emphasizes the relationship between behavioral intentions, self-control, and individual temporal sensitivity in health decision-making. This theory-based intervention has been shown to improve patient self-management behaviors, including adherence to diet and fluids, and reduce therapy-related complications. These results confirm that TST is capable of bridging the gap between intentions and actual behavior, a significant challenge often faced by hemodialysis patients in maintaining long-term treatment regimens.

Compared to other theories already discussed (SDT, HBM, and the 5A Model), TST highlights the unique aspect of self-regulation within the context of time, which is particularly relevant for hemodialysis patients who must adapt to rigorous and repetitive treatment routines. The application of TST broadens the theoretical landscape of hemodialysis education by demonstrating that educational effectiveness depends not only on motivation and risk perception but also on the patient’s ability to regulate their behavior within a specific time frame. These findings underscore the importance of expanding the exploration of various health behavior theories that can be adapted to the hemodialysis context.

These findings emphasize the importance of strengthening theory integration in hemodialysis education design. Studies based on SDT, HBM, and the 5A Model demonstrate broader benefits, not only on knowledge or adherence, but also on psychosocial outcomes and quality of life. Meanwhile, implicit approaches continue to make important contributions as practical innovations, but they should be complemented by a theoretical foundation to make interventions more systematic, measurable, and replicable. Therefore, future research needs to combine the strengths of technological innovation and educational methods with an explicit theoretical framework for health behavior to increase the long-term effectiveness and transferability of interventions across clinical contexts.

## Conclusion

This scoping review shows that educational interventions for hemodialysis patients in the past five years have been dominated by approaches without a formal theoretical framework, focusing on delivery strategies such as teach-back, multimodal education, mHealth, and micro-video. Only a few studies explicitly use health behavior theories as a foundation, namely Self-Determination Theory (SDT), the Health Belief Model (HBM), and the 5A Self-Management Model. These theory-based studies have produced more comprehensive impacts, not only on knowledge and adherence, but also on psychosocial outcomes and patient quality of life. These findings highlight a significant gap in the integration of theory into hemodialysis education practice.

For nursing practice, the results of this review emphasize the need for nurses to adopt theory-based educational interventions to increase long-term effectiveness and strengthen mechanisms for patient behavior change. Educational programs based on SDT, HBM, or the 5A Model can serve as guidelines for developing more structured, measurable, and sustainable interventions. For future research, further exploration of the most effective theories in various cultural contexts and healthcare systems is recommended, as well as long-term evaluation to ensure the sustainability of the intervention’s impact on adherence, self-management, and quality of life of hemodialysis patients.

## Data Availability

All data produced in the present study are available upon reasonable request to the authors

